# Global hotspots and research trends of gut microbiota in sleep disorders: a bibliometric analysis (2011-2024)

**DOI:** 10.1101/2025.08.07.25333265

**Authors:** Li Guo, Changxiao Xie, Xiaoyu Wang, Yujie Xu, Guo Cheng

## Abstract

**Background:** The close connection between gut microbiota and sleep disorders has been confirmed, and research on this topic has increased significantly. However, the overall research trends in this field are unclear. This study aims to assess the status of research on gut microbiota and sleep disorders from a bibliometric analysis perspective and predict future research directions and emerging trends.

**Method:** A systematic search in the Web of Science Core Collection from 2011 to 2024 identified 764 publications. The search terms were “sleep disorders” and “gut microbiota”. Data were analyzed using VOSviewer (version 1.6.17), CiteSpace (version 6.4), Excel (version 16.94), and an online bibliometric platform.

**Results:** From 2011 to 2024, 764 publications on gut microbiota and sleep disorders were identified, showing a steady increase. China (326; 42.67%) was the country with the most publications. The journal Nutrients (42; 5.50%) published the highest number of publications. Zixu Wang (14; h-index = 23) and Shanghai Jiao Tong University (17; 2.23%) were respectively the most productive author and the most prolific institution in this field. The high-frequency keywords were clustered into five thematic areas: cognitive-related topics, psychological and gastrointestinal diseases, metabolic diseases, metabolism and neurodegenerative diseases, and sleep-related topics. Among them, “gut microbiota” (309), “sleep” (133), and “inflammation” (108) were the most researched keywords in the field of gut microbiota and sleep disorders. “Neurodegenerative diseases” and “sleep disorder” were the keywords with the strongest citation bursts from 2022 to 2024.

**Conclusion:** This article provided a bibliometric analysis to systematically summarize the relationship between the gut microbiota and sleep disorders from 2011 to 2024, providing a reference for identifying research trends, key hotspots, and gaps in this field.

## 1. Introduction

Sleep is an important and basic part of keeping the body operating normally [1], such as maintaining health, preventing diseases, and recovering from diseases [2]. However, a significant number of people currently suffer from sleep disorders. It has been reported that 40% to 50% of the global population experience poor or inadequate sleep [3]. Sleep disorders are associated with significant impairments in physical and psychological health and an elevated risk of all-cause mortality [4]. Research has indicated that sleep disorders increase susceptibility to cardiovascular, metabolic, and immune diseases[5], depression [6], and Alzheimer’s disease [7]. Sleep disorders profoundly affect the population, yet their underlying mechanism remains unclear [8]. The latest research showed that sleep disorders were closely related to gut microbiota, which could be used as a prevention and treatment strategy for sleep disorders [9].

Previous studies have demonstrated that acute bacterial infections substantially alter sleep architecture through immune-mediated and neural mechanisms, such as with interleukin-1β-induced increases in non-rapid eye movement sleep [10, 11]. Under dysbiotic conditions, opportunistic pathogens within the gut microbiota may mimic chronic low-grade infections, leading to persistent immune activation and subsequent disruption of sleep homeostasis [12]. The gut microbiota is the most abundant bacterial community in the human body, comprising trillions of bacteria, viruses, archaea, and fungi. It plays an essential role in maintaining human health [13, 14]. The gut microbiota can communicate bidirectionally with the brain via the brain-gut axis pathway and thus interact with sleep patterns [15]. The regulation of the gut microbiota and sleep-wake patterns is mediated via three physiological pathways, including neuronal circuits [16], immune pathway [17, 18], metabolic and endocrine pathways [19, 20]. In recent years, numerous studies have explored the role of gut microbiota in various sleep-related conditions, such as sleep disorders [21]、sleep deprivation [22]、 insomnia [23] and obstructive sleep apnea [24]. With the deepening of research on sleep disorders, the gut microbiota may play a key role in the etiology and pathogenesis of sleep disorders, which deserves further exploration and study [25].

Bibliometrics analysis is a widely used method to identify and summarize research hotspots and new trends in publications within a specific research field or discipline, either quantitatively or qualitatively [26]. Differences from traditional methods, bibliometric analysis provides quantitative data on the distribution of countries, institutions, authors, journals, keywords, and cited literature. In addition, it analyzes correlations among research elements through scientific mapping. In recent years, bibliometric analysis has been extensively utilized in gut microbiota and neuroscience [27, 28].

The literature on gut microbiota and sleep disorders is expanding rapidly but remains scattered [19, 21, 29]. This study utilizes bibliometric analysis to systematically identify research hotspots and development trends in gut microbiota and sleep disorders. It aims to summarize the current findings and assist scholars in understanding the critical research directions in this field.

## 2. Methods

### 2.1 Data origin and search approach

The relevant data were extracted from the Web of Science Core Collection database, covering the period from 2011 to 2024. The search strategy employed is outlined as follows: TS=(gut OR intestin* OR gastrointestine * OR gastro-intestin *OR gut-brain axis) AND TS=(microbiota *OR microbiome OR flora OR microflora OR bacteria) OR TS = (prebiotic OR probiotic OR dysbiosis) AND TS = (sleep disturbance OR sleep deprivation OR sleep disorder OR insomnia OR sleep-related breathing disorders OR central disorders of hypersomnolence OR circadian rhythm sleep-wake disorders OR sleep-related movement disorders OR parasomnias OR sleep apnea syndrome). TS is an acronym for the Topic Search method of the Web of Science database. The search terms were established by identifying relevant subject headings in the Mesh database related to “gastrointestinal”, “microbiota”, and “sleep disorder”, and integrating these with search strategies from prior bibliometric studies on gut microbiota or sleep disorders. Sleep disorders are classified into seven categories globally, including insomnia, breathing-related disturbances, central disorders of hypersomnolence, circadian rhythm irregularities, movement-related issues, parasomnias, and other conditions [30]. The search strategy employed in this study comprehensively covers all the above-mentioned classifications of sleep disorders. Data were systematically retrieved from the included publications, exported in either “plain text format” or “tab-delimited format”, and recorded content as “full record and cited references”. The types of publications included were limited to English literature. The flow chart of the data collection process is presented in Fig 1.

**Fig 1.**
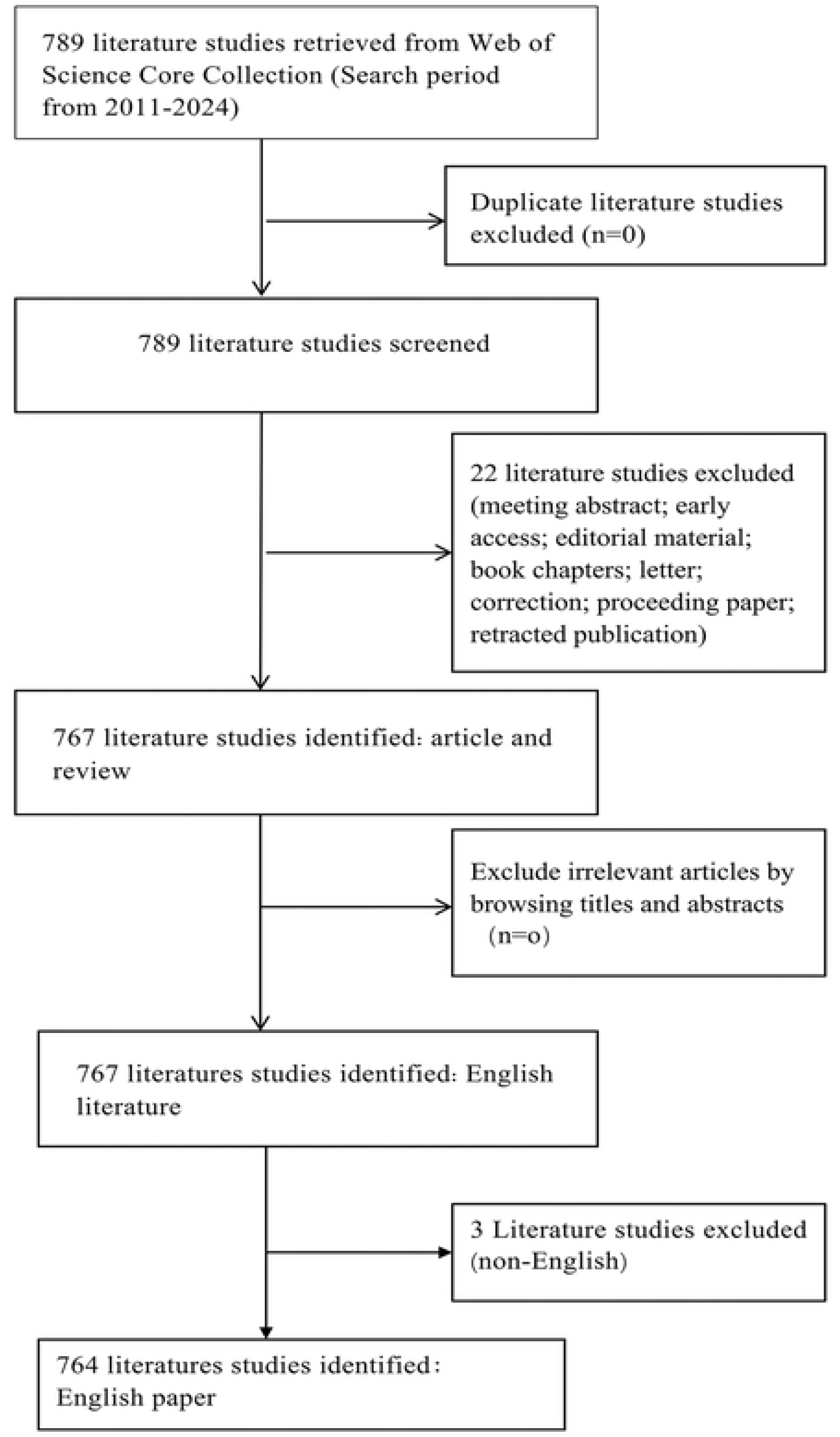
Flowchart for literature inclusion and exclusion.

This study has been registered at https://osf.io/acs5x, and the registered content can be accessed via https://osf.io/k45hc. All original data utilized in this study were obtained from public databases, and ethical approval was not required.

### 2.2 Data analysis

This study adopted a comprehensive analytical approach [31], utilizing VOSviewer (version 1.6.20), CiteSpace (version 6.4), Excel (version 16.94), and an online bibliometric platform to perform integrated quantitative and qualitative analyses of research trends in gut microbiota and sleep disorders [32]. The online bibliometric platform, named Charticulator, is accessible via the following web address: https://donghaoren.org/charticulator/. This study visually analyzes gut microbiota and sleep disorders, focusing on critical aspects, including countries, journals, authors, institutions, keywords, and co-cited references.

This study used a combination of visualization software to analyze the research trends of gut microbiota and sleep disorders. Different visualization tools have distinct focuses and advantages. Compared with a single visualization software analysis, the integrated analysis method can provide more comprehensive and in-depth research in this field. VOSviewer’s advantage lies in the visualization-based analysis of static network structures (co-occurrence network, coupling network, cooperation network) [33]. CiteSpace’s advantage lies in its time series analysis of the evolutionary process of research hotspots. It can provide functions such as research hotspot timeline visualization, burst citation detection, and centrality calculation [34]. Excel can quantitatively analyze the number of publications in the field. The Chartculator online platform can provide a comprehensive and intuitive map of national cooperation networks.

VOSviewer software is widely used for scientific mapping [35]. In this study, VOSviewer was mainly used to analyze the publication counts of journals, the academic output volumes of authors and institutions, and keyword co-occurrence analysis. Keywords co-occurrence analysis was performed using all the keywords in the included publications. Vosview was mainly involved in generating density maps of cited and co-cited journals, the collaboration network maps of authors and institutions, and keywords co-occurrence network analysis. In the figure, nodes represent different research elements, and the size of the nodes reflects the number of publications or the occurrence frequency of keywords. The intensity of the association between nodes can be indicated by the thickness of the lines connecting the nodes or by the total link strength (TLS) [36]. In the different maps, occurrence thresholds for nodes representing various elements were established according to diverse criteria. In the journal citation density map, journals with at least 3 citations are included. In the journal co-citation density map, only journals co-cited at least 260 times are shown. The node thresholds for the author collaboration network and the institutional collaboration network were set such that each author must have published at least 4 publications in this field, and each institution must have published at least 5 publications. The threshold for the keyword co-occurrence network map was set at each word appearing more than 15 times in all the included literature. Nodes that satisfy the threshold requirements will be displayed in the map, and the nodes can be connected to form a network map.

CiteSpace is a widely used visualization tool. This study mainly used it for burst citation detection, co-cited references analysis, and national centrality analysis. CiteSpace generated the top 30 keywords and references with the strongest citation bursts and the top 10 authors and institutions with the strongest citation bursts. In addition, CiteSpace was also used to construct the timeline view map of co-cited references.

The h-index assesses the number of citations to publications issued by authors, and the 2023 Impact Factors (IFs) assesses the average number of citations to publications in journals. CiteSpace computes centrality, a core metric used to measure the importance of nodes (such as countries, literature, authors) in the network structure. Centrality takes values from 0 to 1. The higher the value, the stronger the intermediary role of the node. Nodes with centrality ≥ 0.1 are generally considered to have an important hub role [37]. Hotspots represent cutting-edge or highly cited research themes detected by bibliometric analyses, such as keywords co-occurrence network [38].

## 3. Results

### 3.1 Analysis of annual publications

764 publications met the inclusion and exclusion criteria (Fig 1), including 515 articles and 249 reviews. Since 2019, there has been a rapid increase in publications. Annual publications increased from 126 during 2011–2019 to 638 during 2020–2024, a fivefold increase (Fig 2A). Compared with 2011, the cumulative growth rate of the total number of publications, articles, and reviews in 2020 reached 1200%, 1000% and 1500% respectively (Fig 2B). As shown in Fig 2C, publication trends remained relatively limited and stable until 2016. However, since 2019, research on the gut microbiota in the field of sleep disorders has been continuously increasing. The model fit curve for the growth of the number of publications shows a strong positive correlation with the year of publication (R^2^ = 0.9645, 0.9543, and 0.9409 for the total number of publications, articles, and reviews). We estimated that total annual publications, articles, and reviews will exceed 450, 300, and 100, respectively.

**Fig 2.**
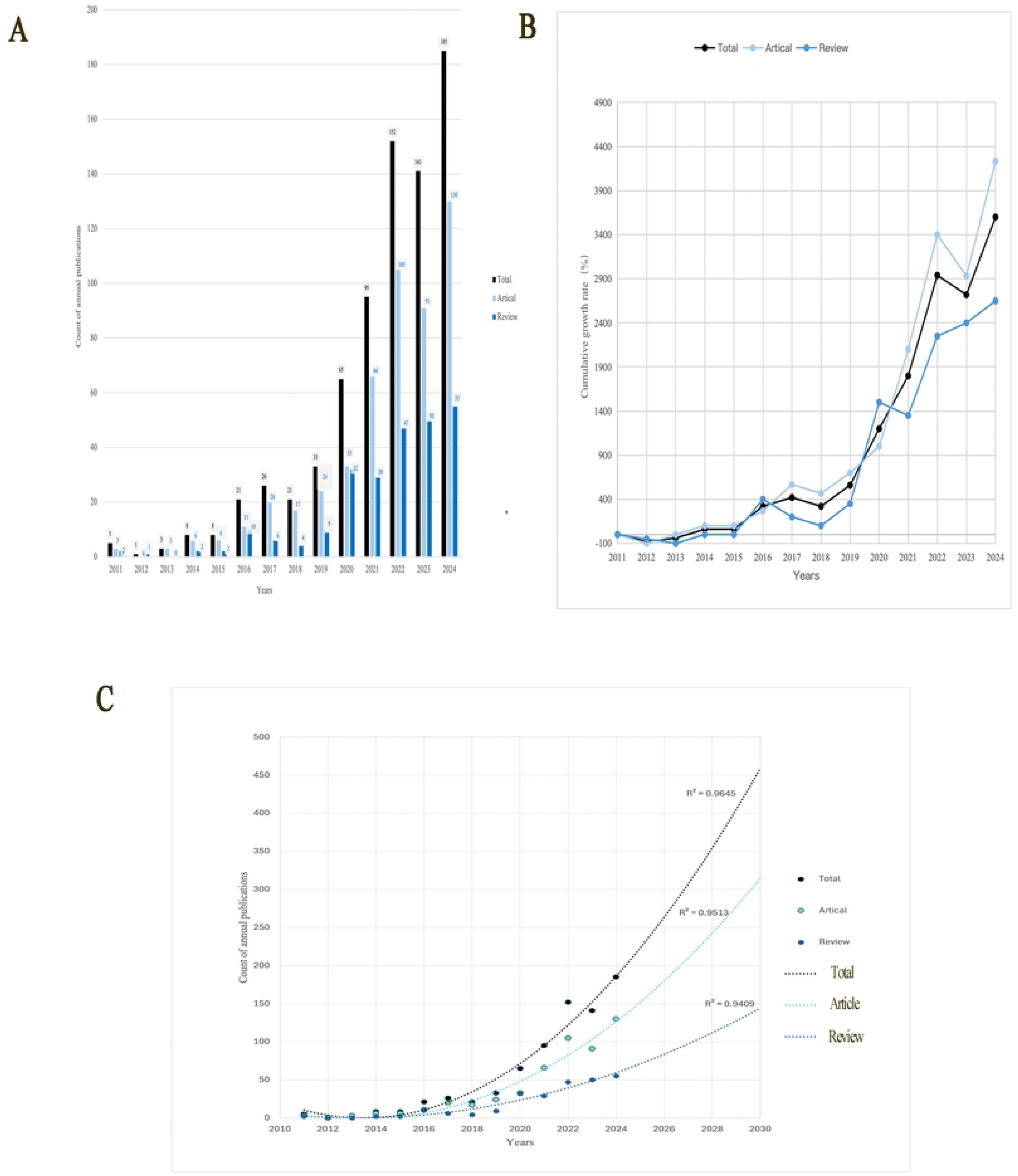
Analysis of the number of publications. (A) Annual publication counts. (B) Cumulative growth rates of publications. (C) Polynomial curve fitting for the growth trend

### 3.2 Analysis of countries

72 countries have contributed to research on gut microbiota and sleep disorders. Table 1 lists the top 10 countries with the highest publication counts and centrality. According to World Bank criteria, among the top 10 countries in the number of publications, except for China and India, which were middle-income countries, the remaining eight were high-income countries [39]. The most publications were in China (326; 42.67%), the United States (161; 21.07%), and Italy (48; 6.28%). Based on centrality measures, the primary core countries in this research field were the United States (Centrality: 0.6), Germany (Centrality: 0.14), and Australia (Centrality: 0.13). Fig 3 shows international cooperation among the top 30 countries with the most publications. The countries with the closest international cooperation were the United States (28 cooperating countries), Australia (26 cooperating countries), and China (21 cooperating countries). The closest cooperation was between China and the United States (TLS: 20), followed by the United States and Australia (TLS: 9), and then the Netherlands and United Kingdom (TLS: 8).

**Fig 3.**
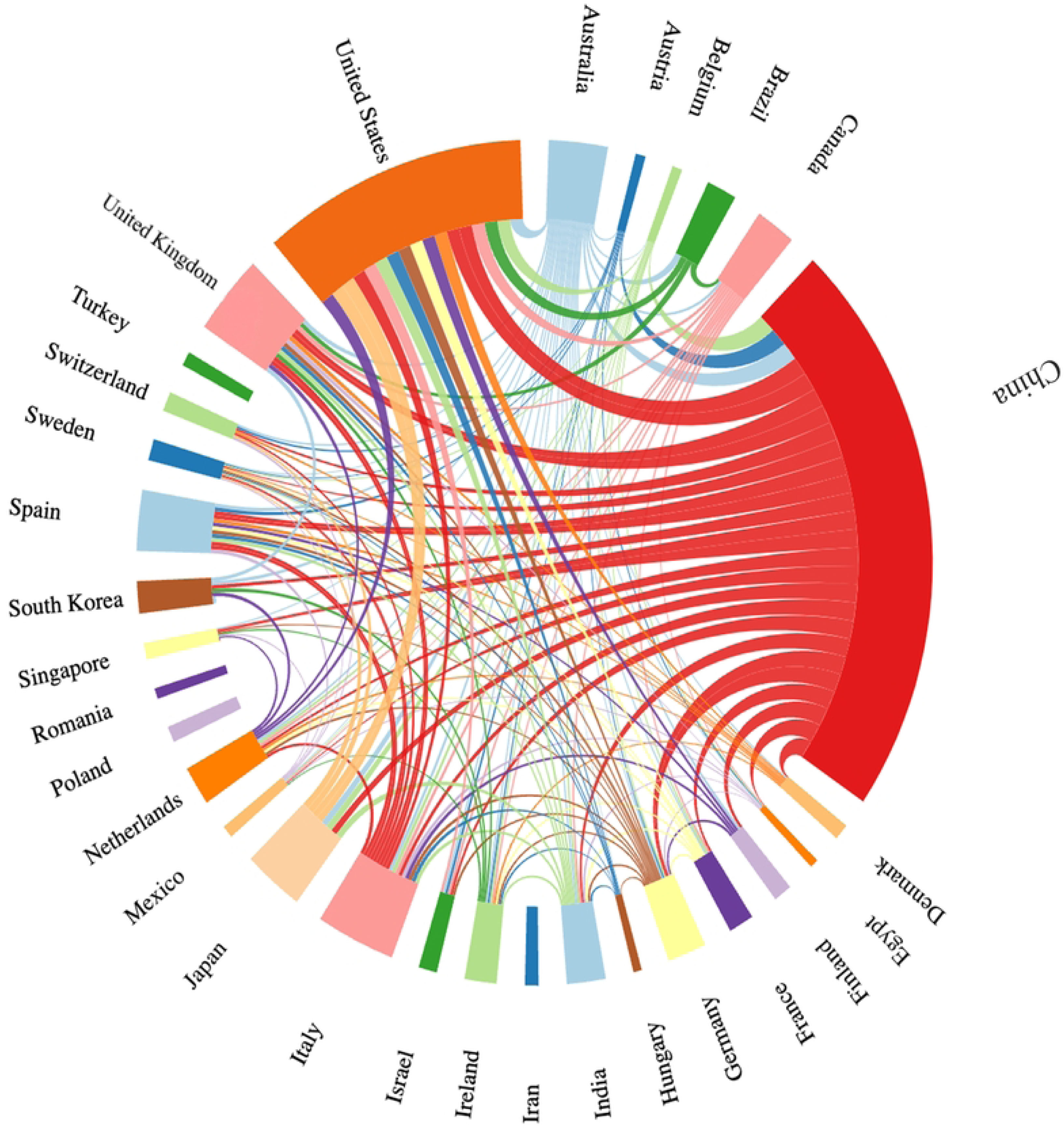
International cooperation among the top 30 countries with the most publications. Different colors indicate different countries; larger color modules correspond to higher publication outputs. Thicker connecting lines represent stronger international collaboration.

**Table 1.**
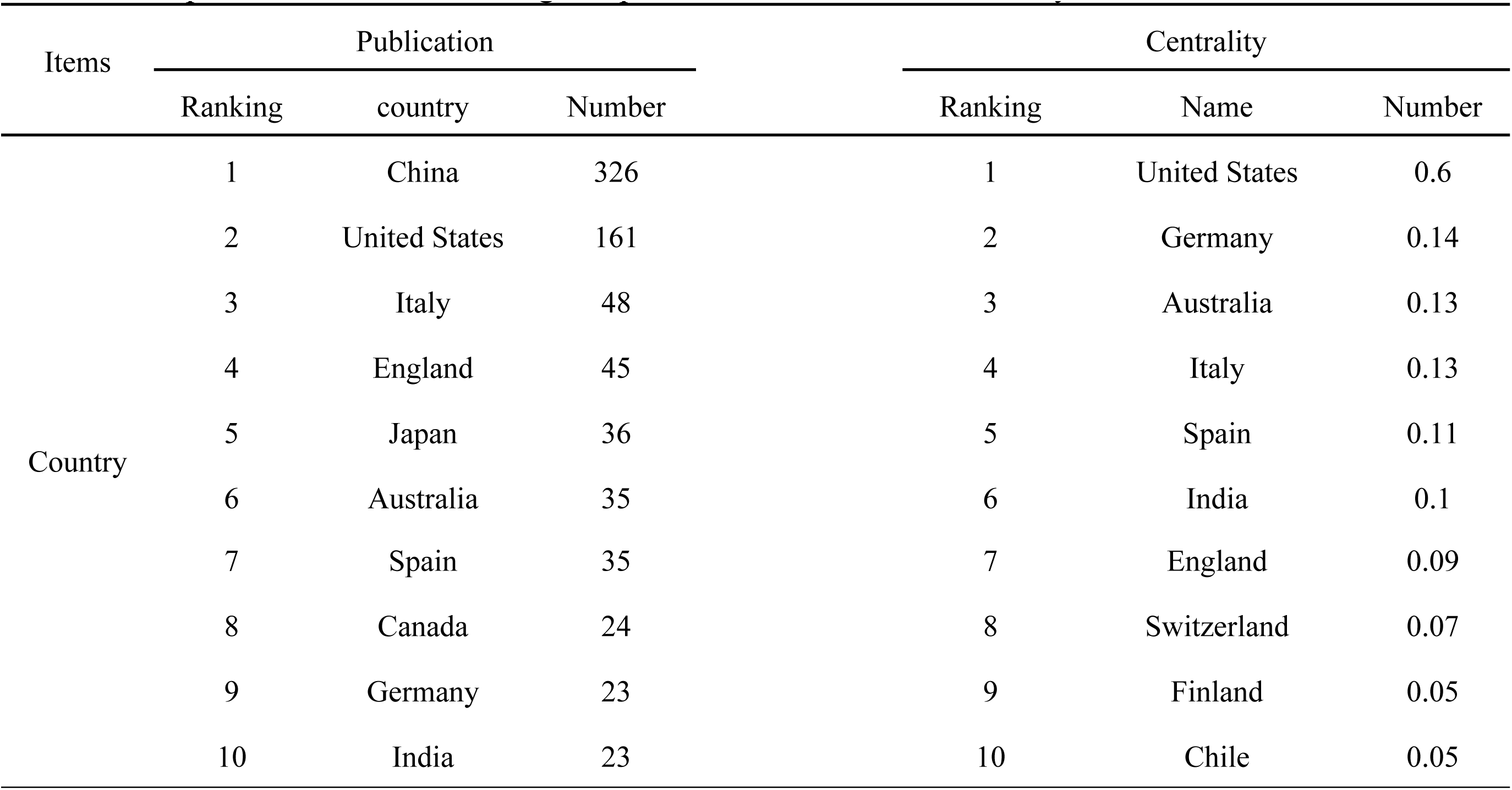
Top 10 countries with the highest publication counts and centrality.

### 3.3 Analysis of journals

764 publications were published in 364 journals. Table 2 lists the top 10 journals with the highest publication counts. Nutrients (42; 5.50%; IF = 4.8) was the most productive journal in the field of gut microbiota and sleep disorders, followed by Frontiers in Microbiology (19; 2.49%; IF = 4) and International Journal of Molecular Sciences (19; 2.49%; IF = 4.9). The 2023 impact factors (IFs) of the top 10 journals ranged from 2.2 to 5.1. According to the Journal Citation Reports (JCR), 50% of these journals were categorized in Q1, while the remaining were in Q2. Fig 4 illustrates the journal density distribution and co-citation density distribution. In the journal density map, Nutrients and Frontiers in Microbiology exhibited a notably high publication output within this research domain. Regarding co-citation density, literature published in PLoS ONE and Nutrients formed a high-frequency co-citation cluster.

**Fig 4.**
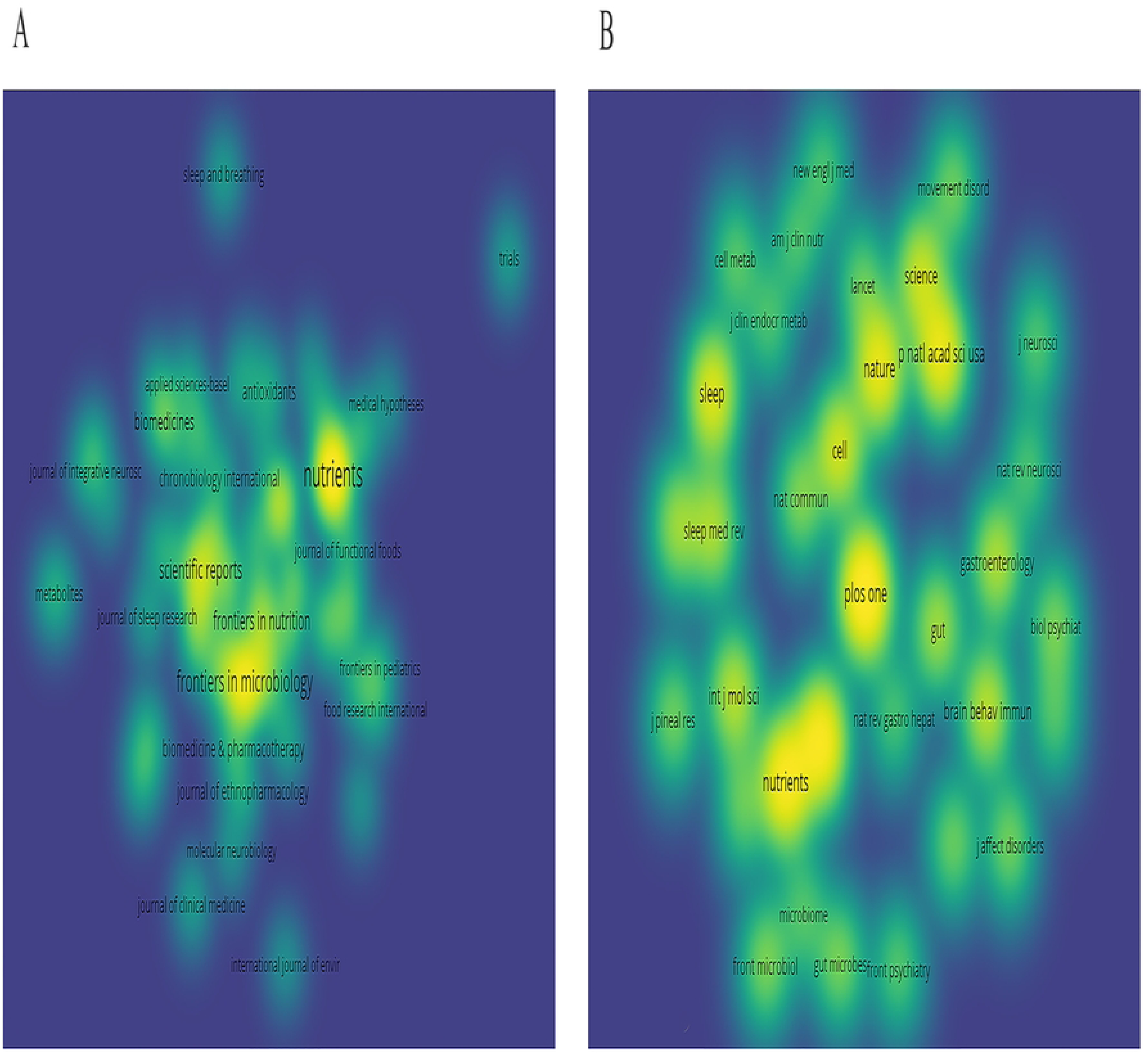
Analysis of journals. (A) Visualization of the density map of cited journals. (B) Visualization of the density map of co-cited journals. Brighter yellow indicates higher journal relevance and attention, whereas blue-green tones suggest lower popularity and less focus.

**Table 2.** Top 10 journals with the highest publication counts.

| Sources | N | Percent | IF (2023) | Category (JCR) |
| --- | --- | --- | --- | --- |
| Nutrients | 4 | 5.50% | 4.8 | Nutrition & Dietetics(O1) |
| Frontiers In Microbiology | 1 | 2.49% | 4 | Microbiology (Q2) |
| International Journal Of Molecular Sciences | 1 | 2.49% | 4.9 | Biochemistry & Molecular Biology |
| Scientific Reports | 1 | 1.70% | 3.8 | Multidisciplinary Sciences (Q1) |
| Frontiers In Psychiatry | 1 | 1.44% | 2.2 | Psychiatry (Q2) |
| Nature And Science Of Sleep | 1 | 1.44% | 3 | Clinical Neurology (Q2) |
| Frontiers In Nutrition | 1 | 1.31% | 4 | Nutrition & Dietetics (Q2) |
| Plos One | 1 | 1.31% | 2.9 | Multidisciplinary Sciences (Q1) |
| Food & Function | 9 | 1.18% | 5.1 | Food Science & Technology |
| Frontiers In Cellular And Infection Microbiology | 9 | 1.18% | 4.6 | Microbiology (Q1) |

### 3.4 Analysis of authors and institutions

4,872 authors from 1,416 institutions contributed to publications on gut microbiota and sleep disorders. Table 3 lists the top 10 most active authors in research on gut microbiota and sleep disorders. The top 3 most influential authors were the top two from China Agricultural University and the third from the University of Missouri. The first was Zixu Wang (14; 1.83%; h-index = 23), with a total of 516 citations. The second was Yaoxing Chen (13; 1.70%; h-index = 28). The third was David Gozal (13; 1.70%; h-index = 108). Fig 5A shows the author collaboration network with 46 authors, TLS of 686, and each author publishing at least four publications. Jing Cao (TLS: 46), Yaoxing Chen (TLS: 46) and Zixu Wang (TLS: 46) had the strongest collaborations with others. Authors in the same cluster, such as Yulan Dong and Zixu Wang showed strong collaboration. In contrast, limited interaction between different clusters suggests restricted cooperation among teams. Besides, Fig 5B shows the top 10 authors with the strongest citation bursts, and the red line segments represent the duration of citations for each reference. Gareau MG and Omahony SM were the most frequently cited authors in recent years.

**Fig 5.**
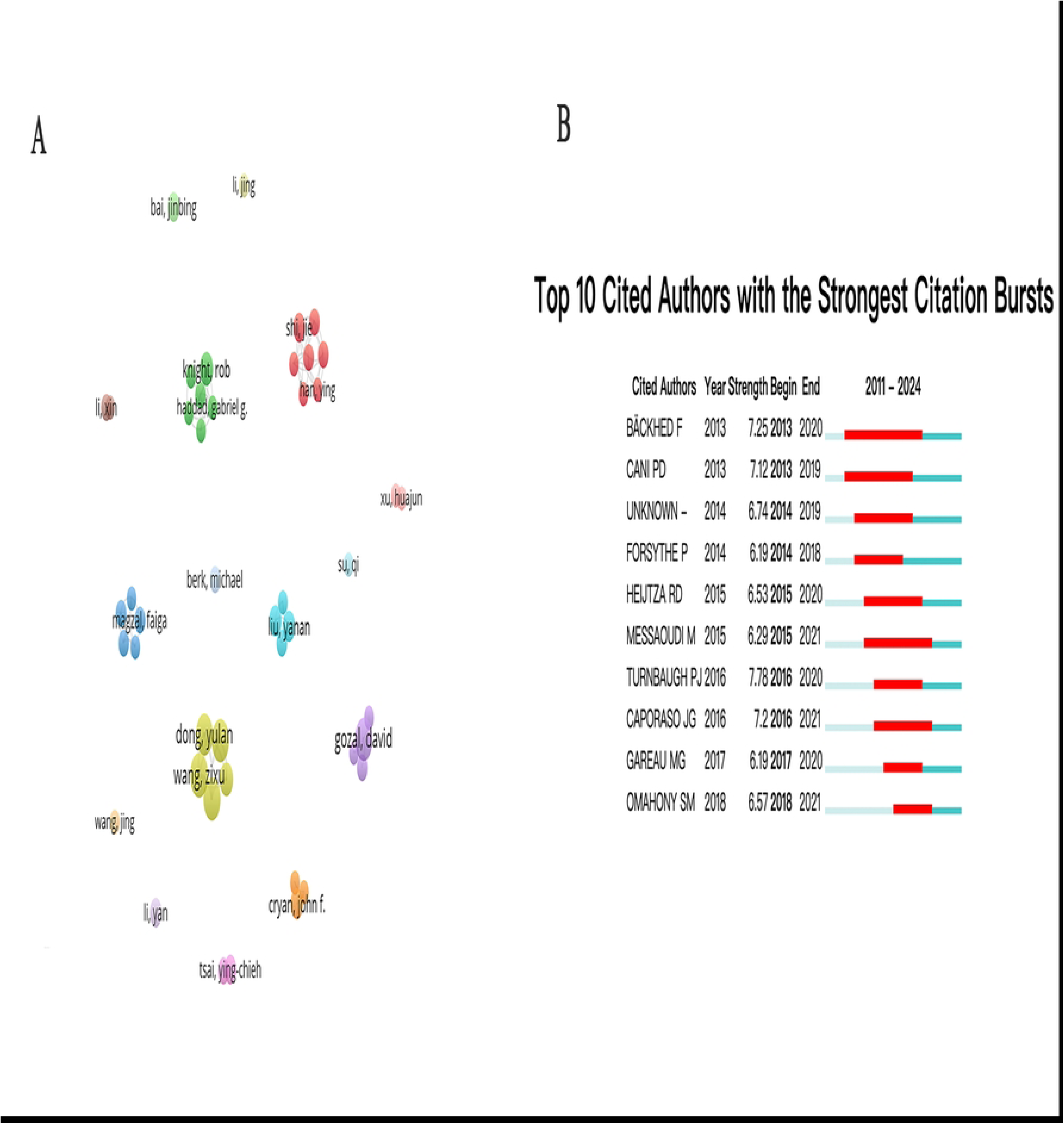
Analysis of authors (A) The collaboration network map of authors. Each node corresponds to an author: larger nodes signify authors with higher publication output. while the links between nodes illustrate collaborative associations among authors. Nodes that exhibit the same color are grouped within the same cluster. (B) Top 10 authors with the strongest citation bursts. Blue bars represent published references, while red bars indicate citation bursts. This rule also applies to Fig 6B, Fig 8 and Fig 9B

**Table 3.** Top 10 authors and institutions with the highest publication counts.

| Ranking | Author<br>(N = 4872) | NP | Total<br>citation | H-index | Institution(N=1416) | NP | Country | Citation |
| --- | --- | --- | --- | --- | --- | --- | --- | --- |
| 1 | Wang, Zixu<br>(China Agricultural<br>University) | 14 | 484 | 23 | Shanghai Jiao Tong<br>University | 17 | China | 292 |
| 2 | Chen,<br>Yaoxing<br>(China agricultural<br>University) | 13 | 482 | 28 | China Agricultural<br>University | 15 | China | 589 |
| 3 | Gozal,<br>David<br>(University of Missouri) | 13 | 946 | 108 | Southern Medical<br>University | 15 | China | 446 |
| 4 | Cao, Jing<br>(China Agricultural<br>University) | 13 | 482 | 13 | University College<br>Cork | 14 | Ireland | 811 |
| 5 | Dong, Yulan<br>(China Agricultural<br>University) | 12 | 477 | 23 | Peking University | 13 | China | 427 |
| 6 | Gao, Ting<br>(China Agricultural<br>University) | 8 | 289 | 30 | Sun Yat-sen<br>University | 11 | China | 401 |
| 7 | Knight, Rob<br>(University of California<br>San Diego) | 8 | 408 | 209 | University of<br>California, San<br>Diego | 11 | United<br>States | 435 |
| 8 | Cryan, John F<br>(University College Cork) | 7 | 853 | 133 | Deakin University | 10 | Australia | 936 |
| 9 | Zhang Xin<br>(Ningbo University) | 7 | 52 | 28 | Ningbo University | 10 | China | 101 |
| 10 | Liu, Yanan<br>(Ningbo University) | 7 | 97 | 21 | Chicago University | 10 | United<br>States | 984 |

NP = Number of publications

Research conducted by 1,416 institutions contributed to 764 publications. Table 3 presents the top 10 institutions with the highest publication counts in this field. Shanghai Jiao Tong University ranked first in terms of publications count in this field (17; 2.23%), followed by China Agricultural University (15; 1.96%) and Southern Medical University (15; 1.96%). Among the top 10 institutions by publication count, the United States has two institutions, but Ireland and Australia each have only one. Notably, the University of Chicago, a leading research institution in the United States, ranked first in citation counts. Fig 6A shows the collaboration network map of institutions. In the map, each of the 58 institutions has published at least 5 publications on this topic. China Agricultural University and Harvard Medical School had the closest cooperation regarding inter-country cooperation. In terms of intra-country cooperation, Chinese domestic institutions were the most frequent. The University of California, San Diego, and Harvard Medical School cooperated closely in the United States. Other institutions had relatively limited collaboration. Fig 6B shows the top 10 institutions with the strongest citation bursts. In the early period, CIBER-Centro de Investigacion Biomedica en Red 2013, CIBERES and the University of Chicago were among the institutions with the strongest citation bursts. In the recent period, Shandong University, Peking University and Southern Medical University had the highest citation counts in this field.

**Fig 6.**
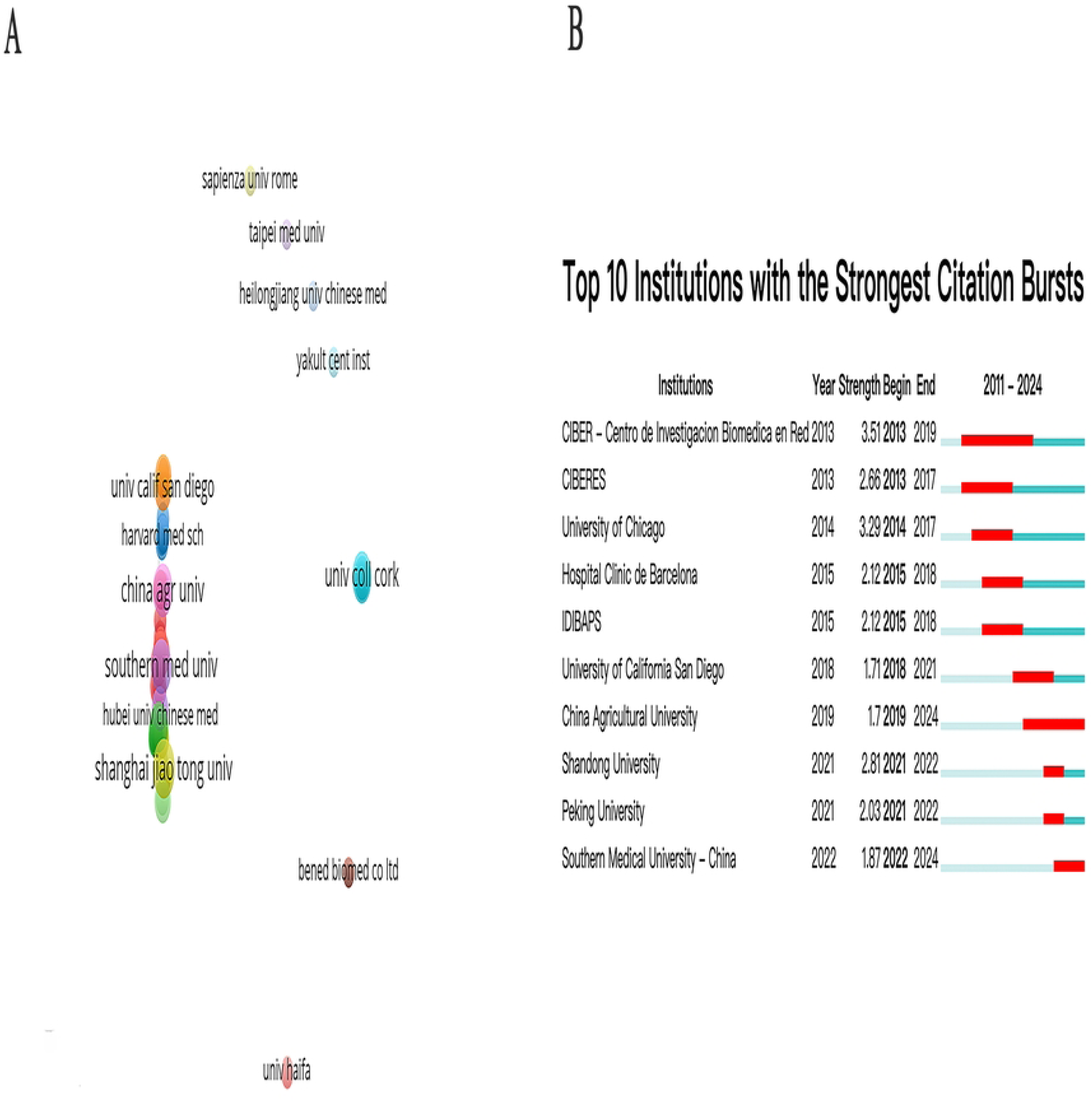
Analysis of institutions. (A) The collaboration network map of institutions. Each node represents an institution: larger nodes signify institutions with higher publication output. (B) Top 10 institutions with the strongest citation bursts.

### 3.5 Analysis of keywords

#### 3.5.1 Keywords co-occurrence network

3,684 keywords were extracted from 764 publications, and 87 keywords appearing over 15 times were analyzed with a TLS of 1,154 (Fig 7). The most frequent keywords were “gut microbiota” (309), “sleep” (133), and “inflammation” (108). The keywords appearing more than 15 times were categorized into five clusters. Cluster 1 (green) mainly refers to sleep disorder and cognitive related topics such as “sleep disorder”, “brain”, “behavior”, “memory”, “expression”. Cluster 2 (red) focuses on psychological and gastrointestinal diseases such as “anxiety”, “depression”, “stress”, “gastrointestinal symptoms”, and “irritable-bowel-syndrome”. Cluster 3 (dark blue) refers to topics related to metabolic diseases such as “hypertension”, “insulin-resistance”, “obesity” and “bloodpressure”. Cluster 4 (yellow) refers to metabolism and neurodegenerative diseases topics, such as “oxidative stress “, “chain fatty–acids”, “gut-brain axis”, and “Parkinson’s disease” topics. Cluster 5 (purple) refers to gut microbiota and sleep topics, such as “gut microbiota”, “sleep”, “insomnia”, and “serotonin”.

**Fig 7.**
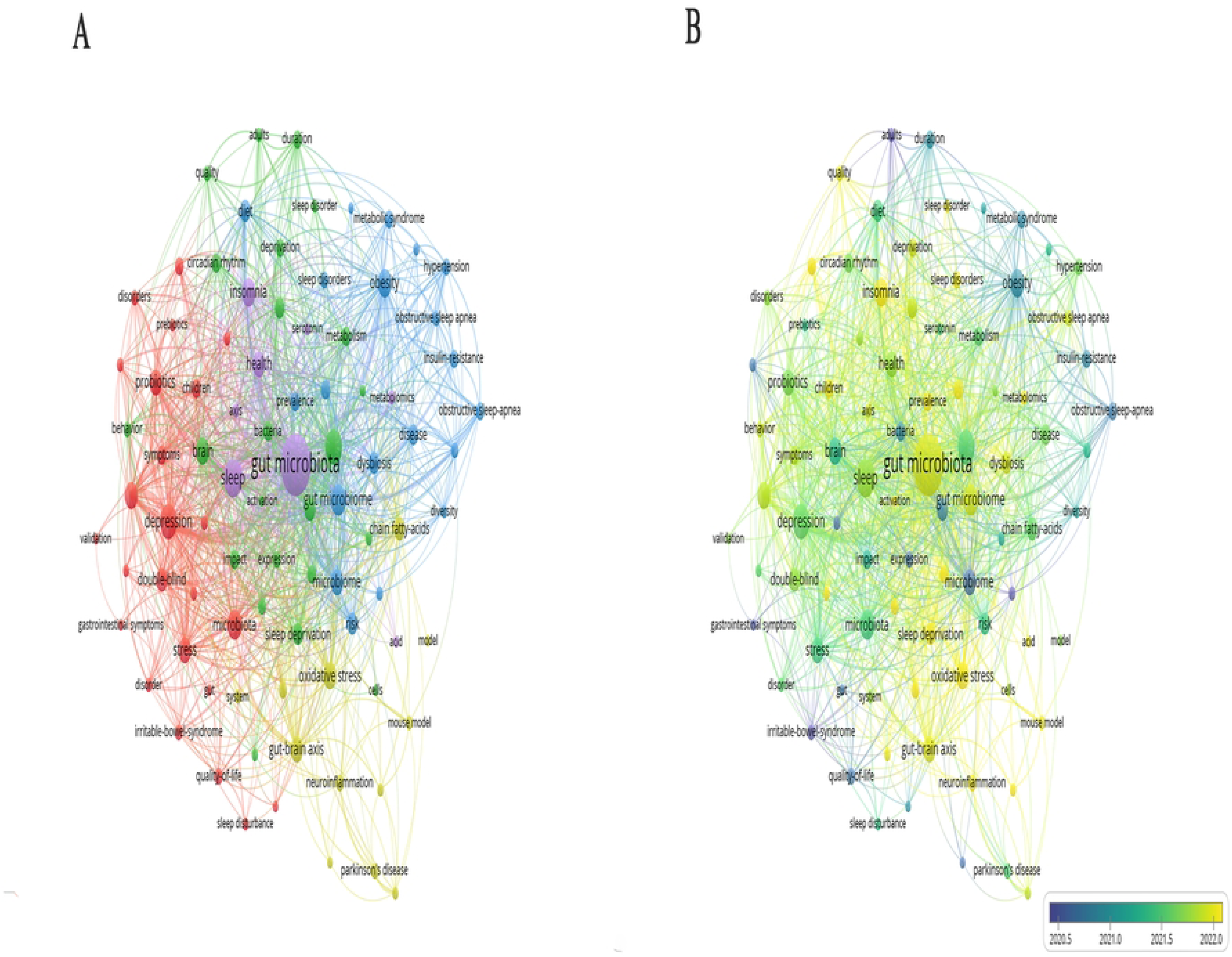
Analysis of the research hotspots on gut microbiota and sleep disorders (A) The network visualization maps of keywords co-occurrence. Keywords are categorized into five categories. Each node corresponds to a specific keyword. Nodes with distinct colors represent different clusters, and the size of a node reflects its frequency of appearance. The lines linking the nodes represent co-occurrence relationships. (B) The network overlay visualization maps of keywords. The overlay maps show the sequence of average publication years with a blueto-yellow gradient.

#### 3.5.2 Keywords with the strongest citation bursts

Fig 8 shows the top 30 keywords with the strongest citation bursts from 2011 to 2024. “Irritable bowel syndrome” and “anxiety-like behavior” received sustained attention from 2012 to 2018. In contrast, “neurodegenerative diseases” and “sleep disorder” were the main research fields from 2022 to 2024.

**Fig 8.**
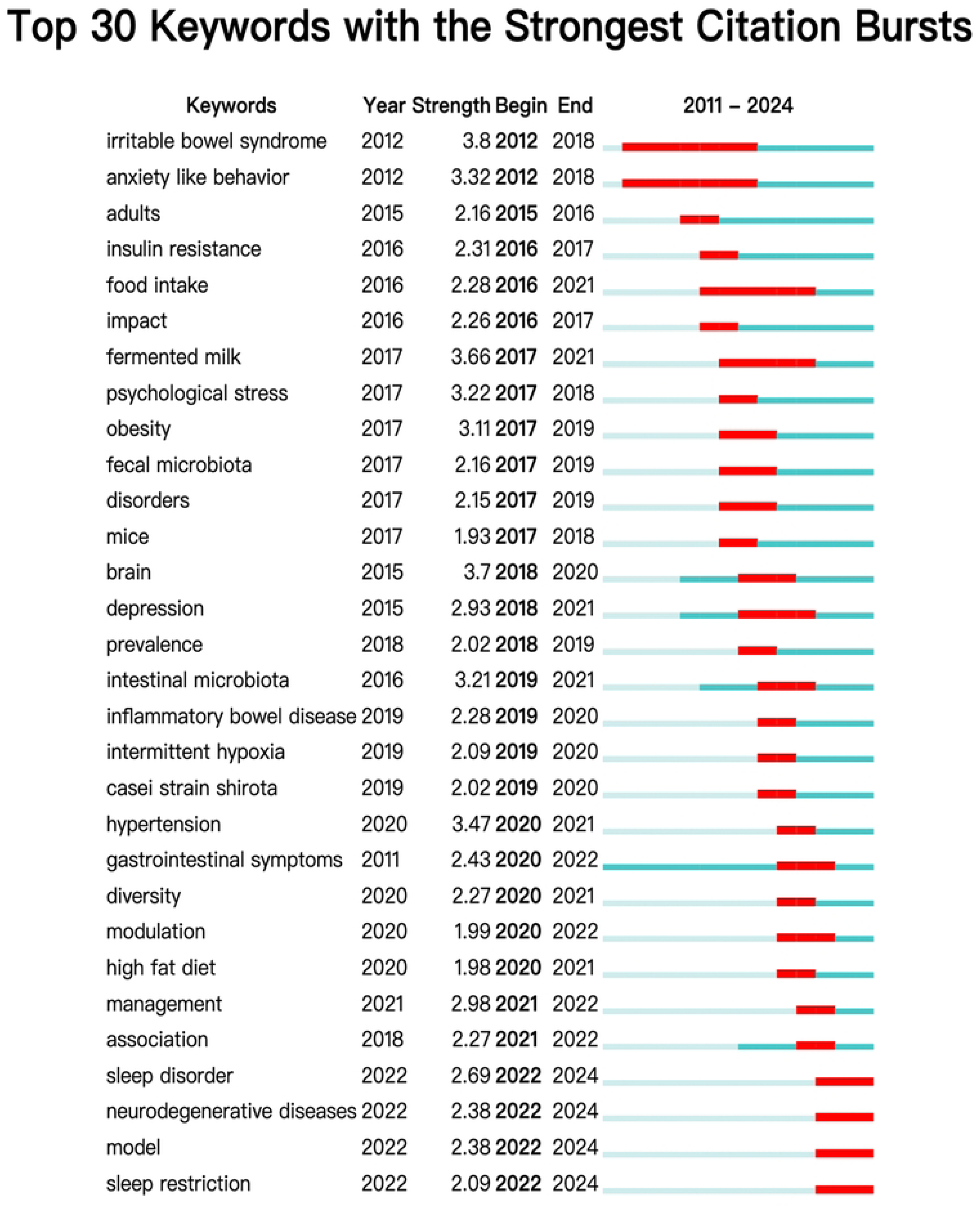
Top 30 keywords with the strongest citation bursts.

### 3.6 Analysis of the references

Table 4 presents the top 10 references with the most citations on gut microbiota and sleep disorders, with citation frequencies ranging from 246 to 918. 8 publications were published in Q1 journals, Galland, L et al. [40] in Q4, and Heintz-Buschart, A et al. [41] in Q2. The most cited publication, by Thaiss, C. A. et al. [42], appeared in Cell (IF=45.5; Q1) and has been cited 918 times. Subsequently, in 2016, Leung, C et al. [43] published “The role of the gut microbiota in NAFLD” in Nature Reviews Gastroenterology & Hepatology (IF=46.4; Q1), cited 705 times. In 2016, Jenkins, F. A. et al. [44] published a study on Nutrients (IF = 4.8; Q1), which has received 543 citations. Among the top 10 cited publications, 7 were reviews, while only 3 were articles.

**Table 4.**
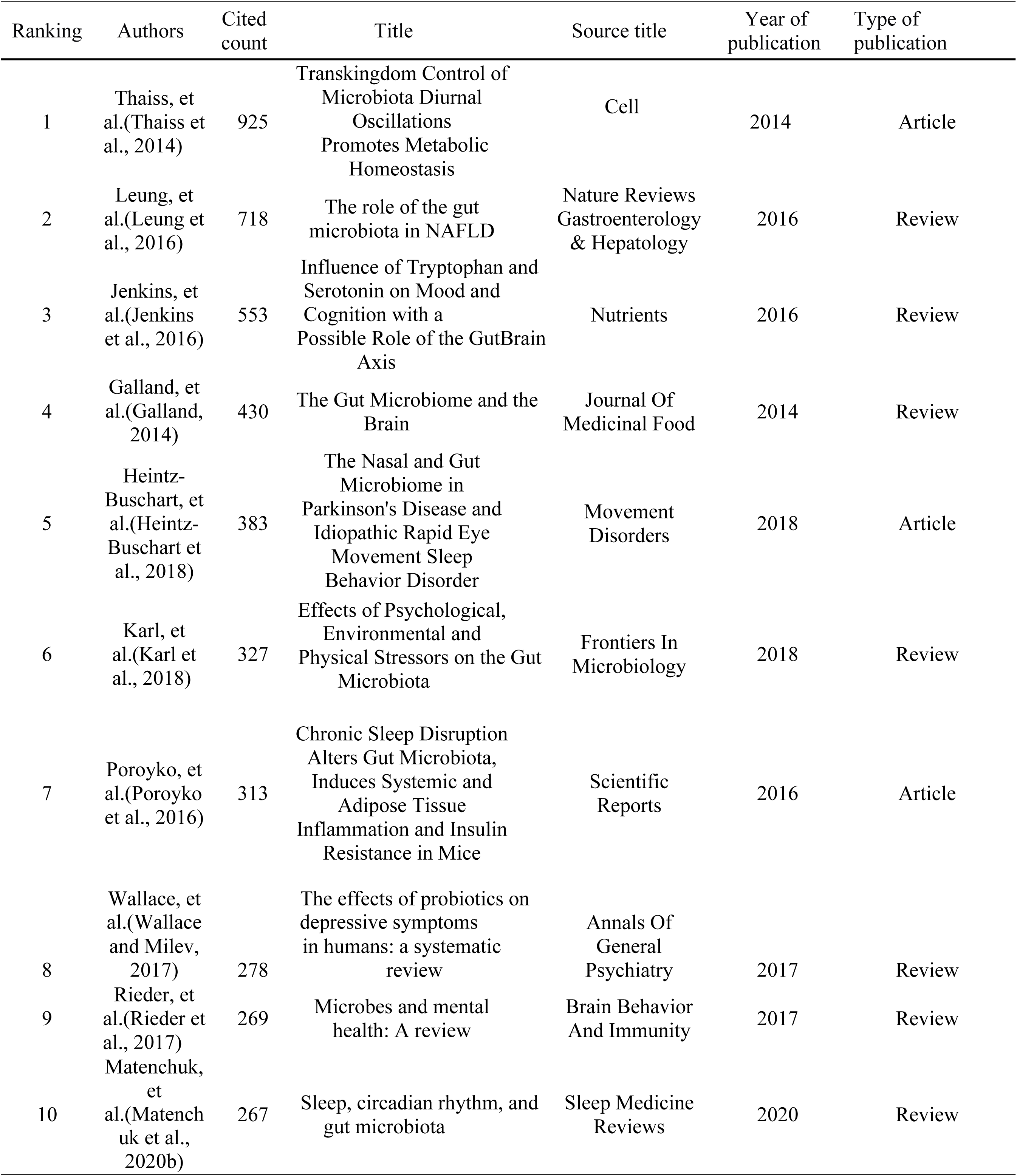
Top 10 references with the most frequent citations.

| Ranking | Authors | Cited count | Title | Source title | Year of publication | Type of publication |
| --- | --- | --- | --- | --- | --- | --- |
| 1 | Thaiss, et al.(Thaiss et al., 2014) | 925 | Transkingdom Control of Microbiota Diurnal Oscillations Promotes Metabolic Homeostasis | Cell | 2014 | Article |
| 2 | Leung, et al.(Leung et al., 2016) | 718 | The role of the gut microbiota in NAFLD | Nature Reviews Gastroenterology & Hepatology | 2016 | Review |
| 3 | Jenkins, et al.(Jenkins et al., 2016) | 553 | Influence of Tryptophan and Serotonin on Mood and Cognition with a Possible Role of the GutBrain Axis | Nutrients | 2016 | Review |
| 4 | Galland, et al.(Galland, 2014) | 430 | The Gut Microbiome and the Brain | Journal Of Medicinal Food | 2014 | Review |
| 5 | Heintz-Buschart, et al.(Heintz-Buschart et al., 2018) | 383 | The Nasal and Gut Microbiome in Parkinson's Disease and Idiopathic Rapid Eye Movement Sleep Behavior Disorder | Movement Disorders | 2018 | Article |
| 6 | Karl, et al.(Karl et al., 2018) | 327 | Effects of Psychological, Environmental and Physical Stressors on the Gut Microbiota | Frontiers In Microbiology | 2018 | Review |
| 7 | Poroyko, et al.(Poroyko et al., 2016) | 313 | Chronic Sleep Disruption Alters Gut Microbiota, Induces Systemic and Adipose Tissue Inflammation and Insulin Resistance in Mice | Scientific Reports | 2016 | Article |
| 8 | Wallace, et al.(Wallace and Milev, 2017) | 278 | The effects of probiotics on depressive symptoms in humans: a systematic review | Annals Of General Psychiatry | 2017 | Review |
| 9 | Rieder, et al.(Rieder et al., 2017) | 269 | Microbes and mental health: A review | Brain Behavior And Immunity | 2017 | Review |
| 10 | Matenchuk, et al.(Matenchuk et al., 2020b) | 267 | Sleep, circadian rhythm, and gut microbiota | Sleep Medicine Reviews | 2020 | Review |

Keywords in the references were visualized and analyzed using CiteSpace. As shown in Fig 9A, they were classified into 12 clusters, including quality of sleep, insomnia, Parkinson’s disease, obstructive sleep apnea, circadian rhythms, dementia with Lewy bodies, microflora, treatment, major depression, nonalcoholic fatty liver disease, lifestyle intervention, and probiotic supplementation. In the timeline view, nodes on the same line represent related research topics in this research area at the corresponding point in time. The size of the point is positively correlated with the research frequency and popularity of the related topic at the corresponding time point. Besides, the cluster groups and nodes on the left side of the line correspond to early research topics, while those on the right side represent recent hot research topics. The topic label on the right of the image corresponds to nodes and clusters on the color lines. Fig 9A shows that early research focused on sleep disorders and cognitive disorders, such as sleep quality, insomnia, dementia with Lewy bodies. It also covered aspects such as treatment and lifestyle interventions. Parkinson’s disease, obstructive sleep apnea, and probiotic supplementation represent recent popular research directions. The references with the strongest citation bursts can indicate the evolution within a knowledge domain [45]. Figure 9B shows the top 30 references with the strongest citation bursts. The first citation burst reference appeared in 2013 [46], and the most recent citation burst reference emerged in 2022 [47, 48]. The reference with the strongest citation bursts (intensity: 24.74) occurred from 2017 to 2021 [49]. Citation bursts for 5 references have persisted until 2024 [48, 50–53]. References experiencing citation bursts between 2016 and 2024 account for 90%.

**Fig 9.**
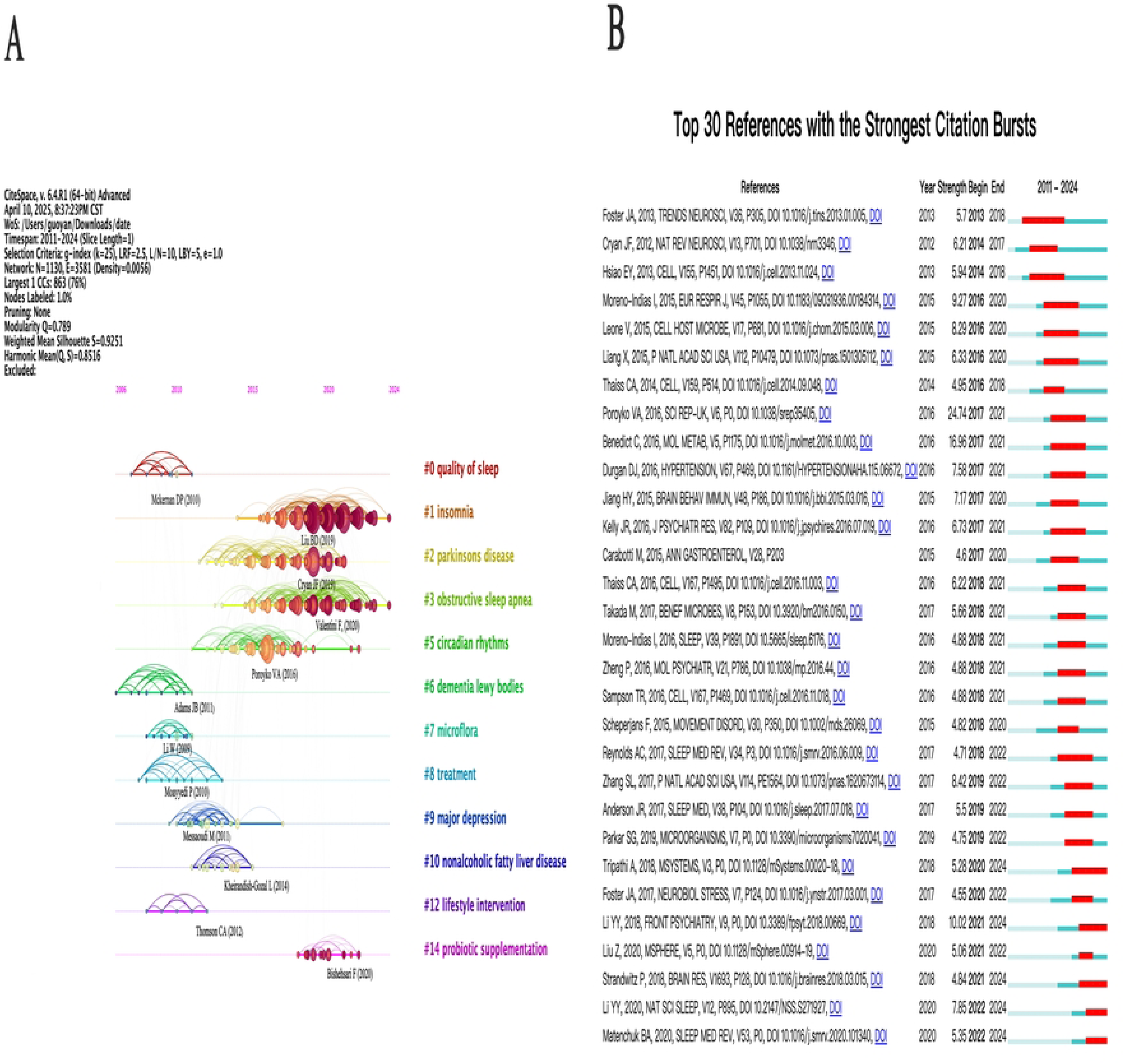
Analysis of co-cited references. (A) The timeline view map of co-cited references. The lines indicate the same cluster. The elements on the right represent the most recent research, reflecting the latest research trends and advancements in this field. (B) Top 30 references with the strongest citation burst.

## 4. Discussion

### 4.1 Analysis of the publication trends

Sleep disorders represent a significant public health challenge in modern society and affect human physical and mental health [12]. Previous studies have shown that insomnia affects 40% of adults and 10% of teenagers. Furthermore, the proportion of the elderly who have difficulty falling asleep is as high as 90% [54, 55], requiring comprehensive research into its root causes and effective treatment options. Gut microbiota is a key research direction in the field of sleep disorders. Abnormal gut microbiota can lead to sleep disorders, and the complex relationship between them highlights the importance of maintaining a balanced microbiota for normal sleep [56]. In this study, the analysis of annual publication results indicated a substantial increase in publications on gut microbiota and sleep disorders. Specifically, the number of publications in this field increased from 168 in 2019 to 638 in 2024, showing a significant growth trend. This indicates that research on gut microbiota and sleep disorders has become a key focus [21]. The sharp increase in publications may be due to projects such as the United States “All of Us” project in 2018 and the 2017 Gut Microbiome Research Program in China, which had advanced research on gut microbiota and sleep disorders [57]. In addition, the surge in sleep disorders during the COVID-19 pandemic has accelerated research in this field [58].

### 4.2 Analysis of national and institutional publication counts

China ranked first in terms of publication count in this field, followed by the United States and Italy. China (326; 42.67%) as the most published country in the field of gut microbiota and sleep disorders, the reason may be that China has many people with sleep disorders. More than 300 million people in China face sleep disorders and related problems [59]. There is an urgent need to explore effective ways to treat sleep disorders. Therefore, China attaches great importance to the development of microorganisms and has allocated substantial financial support to sleep disorders. From 1988-2009 to 2010-2019, the total financing of sleep disorder research in China has increased from more than 10 million yuan to 150 million yuan [60]. Besides, China has launched multiple initiatives, such as the “Brain Science and Brain-like Research” (China Brain Project) and Microbiome Initiative (CMI) [36]. These measures have significantly promoted research on gut microbiota and sleep disorders. The United States ranked second in the world in terms of publication count and collaborated most closely with China in this field. The United States, Germany and Australia have the highest publication centrality in this field. This is because the United States, Australia, and Germany were deeply involved in multiple European research on gut microbiota, such as HBP, which has become the foundation for the research on gut microbiota and sleep disorders [61].

Shanghai Jiao Tong University contributed the most publications in this field. The reason may be its advantages in medical disciplines such as neuroscience and psychiatry at Shanghai Jiao Tong University. However, it only contributed 2.23% of the total publications. Combined with the collaboration network map of institutions, it can be inferred that research institutions in this area were relatively scattered, and no obvious centralized trend has been formed [62]. Among the top 10 authors with the highest publication counts, 50% were from China Agricultural University. Because China Agricultural University may have advantages in specific fields such as animal models, sleep, and metabolism [63]. Chicago University has the highest number of publications cited. The reason may involve three aspects. Firstly, many experts from this institution, such as David Gozal (h-index = 108), and Abdelnaby Khalyfa (h-index = 37). Secondly, this institution started its research in this field earlier than 2014, and its research directions were diverse, involving animal models and human studies [49, 64]. Thirdly, the policies of the American Sleep Research Promotion Program Group have also promoted the development of research in this field at this institution.

### 4.3 Analysis of author and journal publication counts

The collaboration network map of authors reveals that a number of high-productivity authors emerged in the research field of gut microbiota and sleep disorders (such as Yaoxing Chen, Rob Knight, David Gozal), whose research directions covered melatonin, environment, metabolic regulation, and other aspects [65–67]. The research of these authors has greatly advanced the field, especially in terms of sleep disorder mechanisms and clinical applications. However, the author collaborations among different clusters are obviously scattered. This trend limits research breadth and depth. Future development requires strengthening cross-regional and international cooperation to achieve resource-sharing and technology exchange.

Nutrients, Frontiers in Microbiology and International Journal of Molecular Sciences have published the most publications on gut microbiota and sleep disorders. The journal Food &Function was the most influential. In addition, the top 10 journals with the highest publication counts on gut microbiota and sleep disorders accounted for only 20.04% of the total publications, which indicated that the publications were distributed in various journals. This may be attributed to gut microbiota and sleep disorders involved in multidisciplinary fields, including neuroscience, gastroenterology, sleep medicine, microbiology, immunology, endocrinology, and others [21].

### 4.4 Analysis of the most cited references

The most cited references reflect the most valuable and influential discoveries, which can further guide the direction of research in a field. The article “Transkingdom control of microbiota diurnal oscillations promotes metabolic homeostasis” published in CELL [42] was the most cited, with citations rising from 261 in 2019 to 918 in 2024, a 3.5-fold increase. This paper proposed that host circadian rhythms and microbiota circadian disruption can interact through cross-kingdom regulation. It provided new ideas for future exploration of therapeutic strategies for sleep disorders (such as by regulating gut microbiota). The second most-cited article [<u>40</u> focused on the mechanism of gut microbiota in non-alcoholic fatty liver disease (NAFLD). This paper noted that gut microbiota disruption can interfere with metabolism and indirectly affect sleep through inflammatory and immune pathways. It is worth noting that both articles mentioned that probiotics and prebiotics could affect the composition and function of the intestinal microbiota. Thereby indirectly having a positive impact on sleep disorders. Probiotics could be one of the new treatments for sleep disorders, but more clinical studies are needed to verify its effects and determine the optimal dose and strain combination [68]. In addition, among the top 10 most frequently cited references, reviews account for 70%. This phenomenon indicates that sleep disorders and gut microbiota require more original experiments and data to confirm their correlation, which is consistent with the current research trend of microbiota-gut-brain research [69].

### 4.5 Analysis of research hotspots and prospects

Hotspot analysis revealed that gut microbiota and sleep disorders primarily focused on cognitive function and neurodegenerative diseases in recent years, including memory issues, Alzheimer’s disease, and Parkinson’s disease. Additionally, it included psychological disorders (such as depression, stress, anxiety) and metabolism-related disorders (such as hypertension, insulin-resistance, obesity). The “gut microbiota” occupied a central place in the network visualization maps of keywords co-occurrence. It indicated that gut microbiota closely connected with cognitive function, psychological and gastrointestinal diseases, neurodegenerative diseases, metabolic diseases and sleep disorders. It further suggested that modulating gut microbiota may improve related disorders. As the third co-occurrence keyword with the strongest citation bursts, inflammation has been shown to be associated with higher levels of inflammatory markers in individuals with sleep disorders [70]. The underlying mechanism may involve sleep disturbances reducing the number of propionic-producing bacteria, which could result in enhanced intestinal permeability and impaired intestinal integrity. Thus, this process enables the transfer of plasma lipopolysaccharides into the blood, which triggers a systemic inflammatory response [71]. In the future, research can be conducted on the connection between specific intestinal inflammation markers and sleep disorders, providing new methods for the treatment of sleep disorders.

Furthermore, we have found that the evolution of the topic has shifted from the early focus on psychological and physiological disorders (such as irritable bowel syndrome, anxiety-like behavior, hypertension, and insulin resistance) to a more recent concentration on neurodegenerative diseases (such as Alzheimer’s disease, Parkinson’s disease). This shift indicating potential future research frontiers in gut microbiota and sleep disorders. According to the timeline of co-cited references, cognitive disorders such as dementia with Lewy bodies and neurodegenerative diseases such as Parkinson’s disease have been a continuous and hot research area in this field. The most studied types of sleep disorders in recent years were insomnia and obstructive sleep apnea. The latest treatment method was a probiotic supplement. This changing trend highlights emerging research hotspots in the fields of gut microbiota and sleep disorder, offering valuable insights for the treatment of related diseases. High-fat diet and food intake ranked among the top 30 keywords with the strongest citation bursts, which indicates that dietary patterns may influence the regulation of sleep patterns via modulating gut microbiota. Earlier research has shown that consuming a diet high in fat can reduce the alpha and beta diversity of the gut microbiota, increase the Firmicutes/Bacteroidetes ratio, and be negatively associated with sleep disorders [72]. In the future, it may be considered to regulate the gut microbiota by improving the diet structure to treat sleep disorders.

Neurodegenerative diseases are currently a research hotspot, and their association with the gut microbiota and sleep disorders is becoming increasingly apparent. Studies have shown that sleep deprivation can accelerate the aggregation of beta-amyloid (Aβ) and alpha-synuclein (α-syn), which are respectively closely related to Alzheimer’s disease and Parkinson’s disease [73, 74]. Dysbiosis of the gut microbiota may induce metabolic dysregulation, which in turn disrupts sleep quality [75].

However, poor sleep would exacerbate neurodegenerative disease progression through multiple mechanisms such as metabolic dysfunction and systemic inflammation [76]. As neurodegeneration advances, it triggers autonomic nervous system dysfunction, further disrupting gut microbiota homeostasis [77]. This bidirectional interaction could form a vicious cycle of “microbiota-sleep-neurodegeneration.”

### 4.6 Cross-analysis of countries, institutions, keywords

The research on gut microbiota and sleep disorders from 2011 to 2024 has evolved through three stages. In the early stage (2012–2018), studies primarily focused on the interaction between psychological and gastrointestinal factors, with keywords such as “irritable bowel syndrome” and cluster 2 (anxiety-gastrointestinal symptoms). This phase was led mainly by United States-based institutions, including the University of Chicago, which provided foundational experimental evidence for the gut-brain axis. [78]. During the middle stage (2017–2021), the research focus shifted to metabolic disorders, with clusters 3 (obesity, insulin resistance) and 5 (gut microbiota) emerging as key themes. Chinese institutions, such as China Agricultural University, led interdisciplinary investigations into metabolic-gut microbiota-sleep connections, in which keywords such as “fermented milk” mirrored explorations of dietary interventions. [79, 80]. In the most recent stage (2020–2024), attention has increasingly turned to neurodegenerative diseases, marked by the prominence of cluster 1 (sleep disorders) and cluster 4 (Parkinson’s disease, gut-brain axis), as well as the emergence of the term “neurodegenerative diseases.” Chinese institutions, including Peking University, have driven innovation through novel interventions, such as butyrate and melatonin administration [63]. Future research should prioritize the integration of mechanisms connecting metabolic processes and neurodegenerative diseases, thereby facilitating the development of microbiota-targeted precision intervention strategies.

### 4.7 Research limitations

There are some limitations to this study. The types of literature included were limited to articles and reviews, excluding other types of publications. This may lead to the omission of relevant literature. In addition, the scope of our literature inclusion was restricted to English-language publications, which could potentially lead to selection bias. Besides, the data sources of this study were restricted to the Web of Science Core Collection database, which also might lead to the omission of certain relevant studies. However, this is because bibliometric analysis typically recommends using a single database, as it can minimize errors during the data processing phase and enhance the accuracy of bibliometric results [81]. In addition, Web of Science is one of the largest biomedical databases and among the most frequently utilized databases for bibliometric analysis [36]. Therefore, the source of the publications selected in this study is limited to Web of ScienceIn addition. In addition, PubMed, as a professional biomedical abstract database, provides primarily basic publication metadata (such as titles, authors, and abstracts). This represents a significant limitation compared to the Web of Science, particularly in obtaining complete publication records required for comprehensive visualization analyses. Such data constraints may consequently yield incomplete bibliometric visualizations and introduce additional complexities in data processing [82].

## Conclusion

Research on gut microbiota and sleep disorders has developed rapidly, especially in the last five years. China, the United States, and Italy have the most publications on gut microbiota and sleep disorders. The most popular journal was Nutrition. Shanghai Jiao Tong University has the most publications. Zixu Wang was the most active author. David Gozal was the author with the most citations. Research hotspots have transitioned from sleep-related physiological and psychological disorders to cognitive function and neurodegenerative diseases. Obstructive sleep apnea and insomnia are the most studied types of sleep disorders in recent years. In this study, we have three key discoveries in the field of sleep disorders and gut microbiota treatment. Firstly, the gut microbiota plays a central role in this research field and is closely associated with sleep disorders and related diseases, such as Alzheimer’s disease and Parkinson’s disease. This suggests that the gut microbiota may offer novel therapeutic strategies for these diseases. Secondly, our research also found that sleep disorders are closely related to high-fat diets and inflammation. Adjusting the diet structure and treating specific inflammatory markers are potential effective approaches for treating sleep disorders. thirdly, probiotic therapy has become the most popular treatment in the field of sleep disorders and gut microbiota in recent years.

## **7** Author contributions

Li Guo: Writing – original draft, Data curation, Software, Formal analysis.

ChangXiao Xie: Writing – original draft, Methodology, Data curation, Software, Formal analysis.

Xiaoyu Wang: Writing – review and editing, Methodology, Formal analysis. Yujie Xu: review and editing, Project administration, Supervision.

Guo Cheng: Writing – review and editing, Conceptualization, Funding acquisition, Supervision.

## Data Availability

All relevant data are within the manuscript and its Supporting Information files.

## Acknowledgments

We thank Nees. J van Eck and Ludo Waltman for developing VOSviewer and providing it free of charge. We also want to express our appreciation to Professor Chaomei Chen, who developed the CiteSpace software, which is free to use.

## **8** Data availability statement

The original contributions presented in the study are included in the article, further inquiries can be directed to the corresponding author.

## Supporting information

S1 Fig. Fig 1 Flowchart for literature inclusion and exclusion.

S2 Fig. Fig 2 Analysis of the number of publications. (A) Annual publication counts. (B) Cumulative growth rates of publications. (C) Polynomial curve fitting for the growth trend

S3 Fig. Fig 3 International cooperation among the top 30 countries with the most publications. Different colors indicate different countries; larger color modules correspond to higher publication outputs. Thicker connecting lines represent stronger international collaboration.

S4 Fig. Fig 4 Analysis of journals. (A) Visualization of the density map of cited journals. (B) Visualization of the density map of co-cited journals. Brighter yellow indicates higher journal relevance and attention, whereas blue-green tones suggest lower popularity and less focus.

S5 Fig. Fig 5 Analysis of authors (A) The collaboration network map of authors. Each node corresponds to an author: larger nodes signify authors with higher publication output. while the links between nodes illustrate collaborative associations among authors. Nodes that exhibit the same color are grouped within the same cluster. (B) Top 10 authors with the strongest citation bursts. Blue bars represent published references, while red bars indicate citation bursts. This rule also applies to Fig 6B, Fig 8 and Fig 9B

S6 Fig. Fig 6 Analysis of institutions. (A) The collaboration network map of institutions. Each node represents an institution: larger nodes signify institutions with higher publication output. (B) Top 10 institutions with the strongest citation bursts.

S7 Fig. Fig 7 Analysis of the research hotspots on gut microbiota and sleep disorders (A) The network visualization maps of keywords co-occurrence. Keywords are categorized into five categories. Each node corresponds to a specific keyword. Nodes with distinct colors represent different clusters, and the size of a node reflects its frequency of appearance. The lines linking the nodes represent co-occurrence relationships. (B) The network overlay visualization maps of keywords. The overlay maps show the sequence of average publication years with a blueto-yellow gradient.

S8 Fig. Fig 8 Top 30 keywords with the strongest citation bursts.

S9 Fig. Fig 9 Analysis of co-cited references. (A) The timeline view map of co-cited references. The lines indicate the same cluster. The elements on the right represent the most recent research, reflecting the latest research trends and advancements in this field. (B) Top 30 references with the strongest citation burst.

